# Investigating the Amyloid-Tau-Neurodegeneration Framework in Alzheimer’s Disease Using Semi-Supervised Multimodal Imaging Data Fusion

**DOI:** 10.64898/2025.12.11.25341830

**Authors:** You Cheng, Adrián Medina, Cole Korponay, Christian F. Beckmann, David Harper, Lisa Nickerson, the Alzheimer’s Disease Neuroimaging Initiative

## Abstract

**INTRODUCTION:** Alzheimer’s disease (AD) is heterogeneous, complicating diagnosis and prognosis. Uncovering patterns that link abnormalities across amyloid-tau-neurodegeneration (A-T-N) framework may improve prediction of clinical diagnosis.

**METHODS:** We applied SuperBigFLICA (SBF), a semi-supervised multimodal data fusion method, to maps of gray matter density, cortical thickness, pial surface area, amyloid PET, and tau PET in 274 ADNI-3 participants. The model was trained to derive 50 latent components most predictive of a continuous measure of cognitive decline. Latent components’ subject loadings were subsequently used to predict diagnosis (cognitively normal, mild cognitive impairment, dementia) and *APOE4* status using LASSO logistic regression, and were compared against demographic, single-modality, and naïve fusion baselines.

**RESULTS:** While SuperBigFLICA modestly predicted cognitive decline (r = 0.21), SBF loadings-based models outperformed baselines (AUROC = 0.80 for diagnosis; 0.83 for *APOE4*). Amyloid alterations in sensory areas along the sensory–association axis best separated dementia, while a multimodal A-T-N pattern was related to early cognitive decline. Subject loadings on these two patterns were associated with cerebrospinal fluid (CSF) biomarkers, highlighting how CSF AD biomarkers relate to spatial patterns of brain A-T-N burden.

**DISCUSSION:** Semi-supervised multimodal fusion improves prediction and reveals interpretable imaging patterns that predict *APOE4* and clinical diagnoses better than traditional approaches.

## INTRODUCTION

Alzheimer’s disease (AD) is the most common cause of dementia, representing approximately 60–80% of dementia cases and affecting nearly 7.2 million U.S. adults aged 65 and older as of 2025[1]. Clinically, AD is characterized by a progression of changes with aging that begin from cognitively normal (CN) status through mild cognitive impairment (MCI) to Alzheimer’s dementia. Significant clinical and neuropathological heterogeneity across these stages complicates early diagnosis and targeted therapeutic interventions[2].

Multimodal neuroimaging provides a rich and comprehensive view of brain changes in AD, as different imaging modalities are sensitive to both overlapping and distinct neurobiological processes. For example, structural magnetic resonance imaging (MRI) provides a view of neurodegeneration, positron emission tomography (PET) of molecular pathology, and functional MRI (fMRI) of brain circuitry integrity. Combining structural MRI with amyloid⍰PET markedly improves diagnostic sensitivity and prognostic accuracy compared with either modality alone[3,4]. In addition to disease signals that are modality-specific, some neurodegenerative processes are convergent across modalities, with changes in the underlying processes manifesting across multiple modalities. For example, brain atrophy observed on MRI is highly correlated with tau pathology detected via tau-PET, particularly in medial temporal regions[5]. Capturing both modality⍰unique and shared signals is therefore essential for a more comprehensive mechanistic model. Furthermore, AD pathology manifests spatially distinct patterns across modalities. For example, high early amyloid burden but minimal atrophy may be observed in regions such as the frontal cortex, conversely significant hippocampal atrophy may be observed with relatively low local amyloid deposition[6].

Despite advances in machine learning models for predicting clinical diagnoses or AD risk factors (e.g., *APOE4* carrier status), existing approaches predominantly focus on categorical predictions (e.g., MCI to AD conversion) or rely on limited non-imaging and/or imaging modalities (e.g., only neuropsychological tests[7–10], diffusion tensor imaging [DTI][8], MRI[9–15], fMRI[14,16], electroencephalogram [EEG][17]). These models typically lack interpretability regarding how features across modalities collectively relate to the target variable, limiting insight into underlying disease mechanisms. Moreover, traditional pairwise modality comparisons, such as voxel-based analyses between fluorodeoxyglucose (FDG) PET hypometabolism and MRI atrophy, cannot scale to incorporate broader multimodal data[18]. This represents a critical gap in the literature, as leveraging shared variance across imaging modalities toward target variable prediction may yield more biologically meaningful early detection of heterogenous disease.

Addressing these limitations, recent advances in multimodal neuroimaging fusion enable the identification of multimodal brain patterns via joint analysis that effectively increases both the power of detecting convergent effects and the interpretability of the findings[19]. These patterns are typically linked post hoc to behavioral, clinical and demographic phenotypes[20– 22]. In this study, we apply a modified version of FMRIB’s Linked Independent Component Analysis (FLICA), a multimodal Bayesian independent component analysis method for data fusion, that has been scaled for big data and modified for simultaneous supervised learning of a target variable, i.e. SuperBigFLICA or SBF[23]. SBF is a cutting-edge data fusion approach that capitalizes on the benefits of multiple modalities being measured within each subject while modeling shared covariance across imaging modalities to identify linked multimodal latent spatial components that are maximally predictive of a continuous target variable. This simultaneous unsupervised multimodal decomposition with supervised target prediction enables both robust prediction and insights into the brain spatial patterns most associated with these outcomes. This approach bridges the gap between prediction and neurobiological understanding and increase [23]sensitivity to gradual disease progression and individual variability, allowing subtler brain–behavior relationships to emerge.

To demonstrate the utility of SuperBigFLICA (SBF) for multimodal imaging in AD, we trained the model on cognitive decline (clinical dementia rating – sum of box or CDR-SOB), a continuous measure of cognitive status that is closely aligned with clinical diagnosis, using it as a proxy training variable. We hypothesized that (i) cross-modal covariance patterns spanning amyloid, tau, and neurodegeneration (A-T-N) modalities would capture variance relevant to clinical outcomes and improve prediction of clinical diagnoses, and (ii) latent loadings derived from these patterns could be further leveraged to enhance prediction of genetic risk, specifically apolipoprotein E ε4 (*APOE4*) carrier status.

## 2 METHODS

### 2.1 Study participants

Data used in the preparation of this article were obtained from the Alzheimer’s Disease Neuroimaging Initiative (ADNI) database (adni.loni.usc.edu). The ADNI was launched in 2003 as a public-private partnership, led by Principal Investigator Michael W. Weiner, MD. The primary goal of ADNI has been to test whether longitudinal MRI, PET, other biological markers, and clinical and neuropsychological assessment can be used to track the progression of MCI and early AD.

Data were obtained from ADNI⍰3, which includes tau-PET. We selected one visit per participant that contained complete data for (1) Aβ⍰PET, (2) tau⍰PET, (3) 3T T1⍰weighted MRI, (4) demographics (age, sex), and (5) Clinical Dementia Rating–Sum of Boxes[24] (CDR⍰SOB), which sums six domains (memory, orientation, judgment/problem solving, community affairs, home/hobbies, personal care) each scored 0–3 for a total of 0–18. One proposed translation [25] interprets scores of 0 as cognitively normal, 0.5–4 as MCI/subjective cognitive decline (SCC), 4.5–9 as mild dementia, 9.5–15.5 as moderate dementia, and 16–18 as severe dementia. The resulting cohort spanned CN, MCI, and AD diagnoses; amyloid status was not used as an inclusion criterion so that the sample would represent cognitive aging broadly rather than the biomarker⍰defined Alzheimer’s continuum. All ADNI⍰3 enrollees satisfy a common set of eligibility rules[26]. Briefly, participants are 55–90⍰years old, have a Geriatric Depression Scale (GDS) score <⍰6, at least a sixth⍰grade education (or equivalent work history), and adequate vision and hearing for neuropsychological testing. Exclusion criteria include major neurologic disorders other than suspected AD (e.g., Parkinson’s disease, hydrocephalus, or significant head trauma), MRI contraindications, recent major psychiatric illness or substance abuse, clinically significant systemic disease, residence in a skilled⍰nursing facility, and use of prohibited psychoactive or anticoagulant medications. Cerebrospinal fluid (CSF) samples, when available, were included if collected within one year of the scan were assessed with Roche’s Elecsys assays.

### 2.2 Image acquisition

In ADNI-3, imaging followed standardized protocols to ensure consistency across sites[26]. Structural MRI used 3T scanners with a 3D T1-weighted MPRAGE sequence[27]. Amyloid PET employed [^18^F]florbetapir (FBP) or [^18^F]florbetaben (FBB), with acquisitions starting 50 minutes post-injection for FBP and 90 minutes for FBB (four 5-minute frames). Tau PET used [^18^F]AV-1451 (flortaucipir [FTP]), acquired in six 5-minute frames starting 75 minutes post-injection[28].

### 2.3 Image preprocessing

Structural image data preprocessing was performed using fMRIPrep 20.2.7[29,30][Supplementary Material 2], which is based on Nipype 1.7.0[31,32] (see Appendix I for details). Pre-processed T1-weighted images were used to compute probabilistic gray⍰matter maps (GM) using FSL-VBM[33–36], masked to retain only gray-matter regions, as well as cortical surface maps of cortical thickness (CT) and pial surface area (PSA) using FreeSurfer[37] for each participant.

For all PET image data, we used preprocessed single-frame Standardized Uptake Value Ratio (SUVR) images downloaded from the LONI repository (https://ida.loni.usc.edu), which were already motion-corrected, frame-averaged, co-registered to each participant’s T1-weighted MRI, intensity-normalized to a cerebellar reference region, and nonlinearly warped to MNI152 space with 2⍰mm isotropic resolution[38–40]. To harmonize FBP and FBB SUVR maps, we converted voxel-wise SUVR values to Centiloid units using ADNI-provided transformation equations[41]. Amyloid PET data were further masked to gray-matter voxels to minimize nonspecific white-matter signal, while tau PET data were masked to gray matter with exclusion of white matter and cerebellum.

### 2.4 SuperBigFLICA model

We used SuperBigFLICA (SBF) [23], a semi-supervised multimodal fusion framework, to jointly decompose neuroimaging data and predict clinical decline (see details and open access code at https://github.com/ANSR-laboratory/SuperBigFLICA_McL). Five voxel-/vertex-wise maps per participant (GM, CT, PSA, amyloid-PET Centiloid [AMY], tau-PET SUVR [TAU]) were each concatenated across subjects to create five modality series that were fed into SBF to decompose the data into 50 latent components that were maximally predictive of CDR-SOB (see Figure 1). For SBF’s embedded cross-validation, data were randomly partitioned into training⍰(70⍰%), validation⍰ (15⍰%), and test⍰ (15⍰%) sets, with all participants from a given imaging site assigned to the same split to prevent site⍰related data leakage. The analyses were conducted in Python 3.9 using PyTorch 2.1.2.

**Figure 1.**
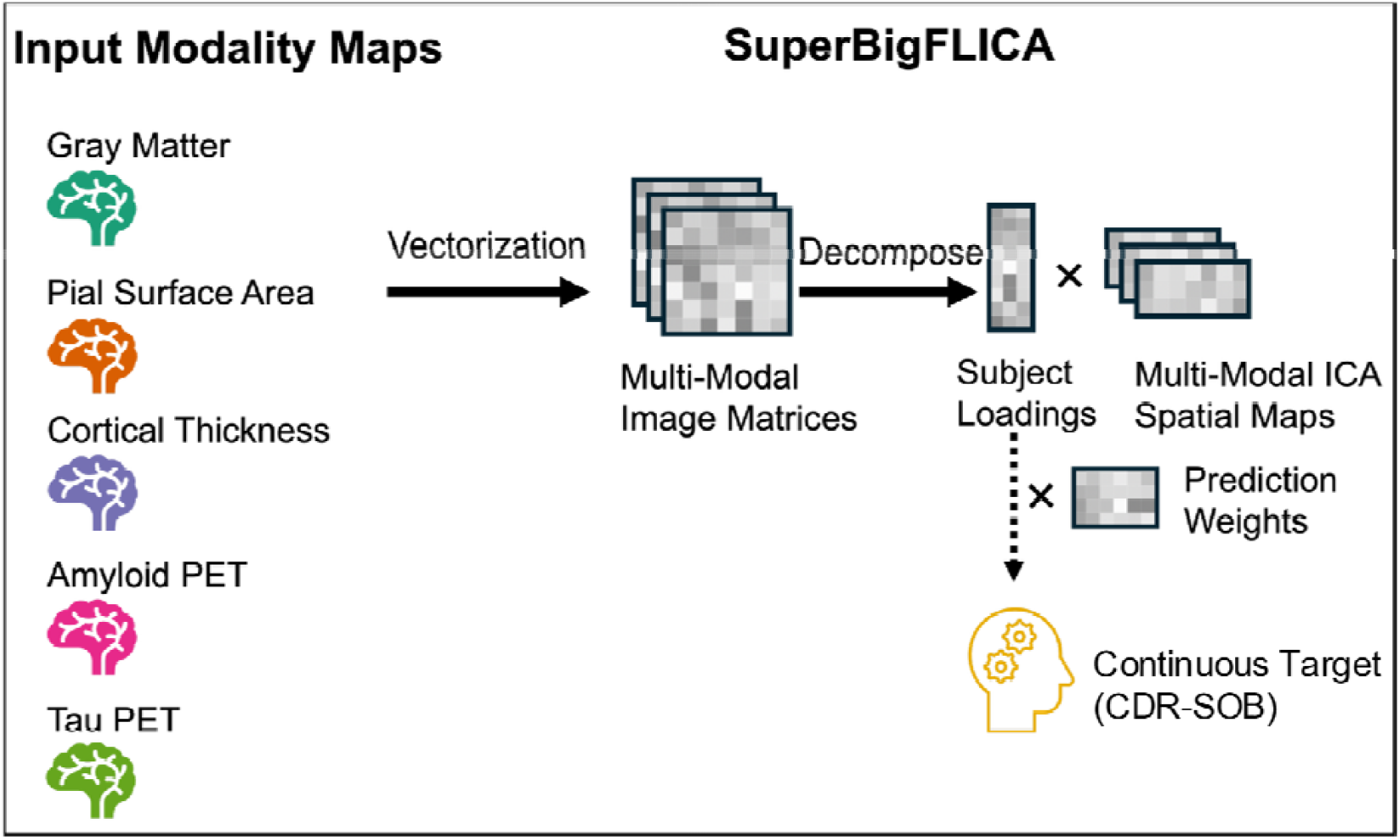
Implementation of the SuperBigFLICA approach for semi-supervised multimodal fusion and phenotype discovery. Workflow of the SuperBigFLICA approach in this study. Inputs include five imaging modality maps from the ADNI-3 dataset: gray matter density (GM), cortical thickness (CT), pial surface area (PSA), amyloid PET Centiloid (AMY), and tau PET SUVR (TAU). These maps, together with the continuous target (i.e., CDR-SOB scores), were fed into the SuperBigFLICA algorithm for multimodal learning. The outputs are the subject loadings, multimodal spatial maps and their prediction weights for maximal prediction of the target variable.

The model produced (i) participant-level latent loadings for both training and test sets, (ii) spatial maps for each of the 50 components, and (iii) outcome predictions evaluated by the correlation between predicted and observed scores in the independent test set. The output spatial maps were thresholded individually using mixture model–based thresholding in FSL MELODIC (mmthresh = 0.5), which explicitly models the distributions of the noise and signal classes in each map.

### 2.5 Prediction of clinical diagnoses and *APOE4* carrier status

The SBF model is designed to identify latent representations that generalize across related outcomes[23], therefore, we tested whether subject loadings derived from the SBF model trained to predict CDR-SOB could transfer to clinical diagnoses and *APOE4* carrier status. Specifically, binary Least Absolute Shrinkage and Selection Operator (LASSO) logistic regression models were trained to classify both clinical diagnosis (CN, MCI, dementia) and *APOE4* carrier status. The primary input feature set comprised latent loadings from all 50 SBF components. LASSO was chosen to prevent multicollinearity while enabling feature selection. Model fitting used the latent loadings from the SBF training set, and evaluation used the latent loadings from the SBF test set.

Prediction performance of SBF loadings was compared against additional baseline feature sets including: (1) demographics (age, sex, education); (2) single-modality imaging features derived from the top 50 principal components (PCs) of each raw modality—GM, CT, PSA, AMY and TAU; (3) a multimodal feature set composed of the top 10 PCs from each modality (50 total); (4) a simple “naïve” fusion approach in which PCA was applied to all modalities concatenated together, from which the top 50 PCs were extracted; and (5) independent component analysis (ICA) analogs of (2–4), in which 50 independent components were extracted in parallel to PCA baselines. PCA was implemented using IncrementalPCA in Python 3.11.7 scikit-learn v 1.7.2, while ICA baselines were generated with FastICA in scikit-learn.

For training the SBF loadings-based model and all baselines, logistic regressions were fit with L1 (LASSO) regularization using the caret v7.0.1 interface to glmnet v4.1.9 package in R v4.3.2, tuning λ across a log-spaced grid from 1×10^−3^ to 1×10^1^. Cross-validation was performed in a site-wise manner: participants from the same acquisition site were kept together, with 5 folds ensuring ~80% of sites for training and ~20% for validation in each fold, with features z-scored within each training fold. Performance on the independent test set was evaluated with the Area Under Receiver Operating Characteristic curve (AUROC; pROC package v1.18.5) and the Area Under the Precision–Recall (PR) Curve (AUPRC; PRROC package v1.4) in R v4.3.2, with 2,000 bootstrap confidence intervals. Class imbalance was handled using inverse-frequency sample weights (assigning half the total weight to the rarer class). Due to the limited size of the independent test set, threshold-dependent metrics (accuracy, sensitivity, specificity, precision, recall, F1 score, balanced accuracy) were derived from cross-validated predictions within the training set, using the threshold that maximized balanced accuracy. Cross-validated metrics are reported because the SBF loadings are derived from the final trained SBF model, making it infeasible to recompute SBF within each fold without severe instability; thus CV reflects model consistency under the learned latent representation, while the small, fully independent test set provides an unbiased (but less stable) estimate. For multi-class outcomes (i.e., clinical diagnosis), one-versus-one (OvO) classification was conducted, and macro-averaged scores across all pairwise comparisons were reported.

Additionally, loadings from the components most predictive of diagnoses and *APOE4* were correlated with CSF amyloid (Aβ 42) and tau (phosphorylated tau or pTau 181) in the subset of participants with available CSF data to help interpret and validate their neuropathological associations—including for components derived from modalities where links to amyloid and tau are not direct. Only CSF samples collected within one year of the imaging visit were included, and when multiple samples were available, the draw closest to the imaging date was chosen.

## 3 RESULTS

### 3.1 Study participants

The ADNI-3 sample included 274 participants with all required data split into training (n = 192), validation (n = 41), and test (n = 41) sets. The mean age was 70.8 years (SD = 6.9), and 56.2% (154/274) were female. Most were White (92.7%) and not Hispanic/Latino (96%). Overall, educational attainment was high: 44.2% held a graduate degree, 46% had some college or a college degree, and 9.9% had a high school education or less. *APOE4* carriers comprised 40.5% (111/274). Clinically, 63.9% were cognitively normal (CN), 21.9% had amnestic MCI, and 14.2% had dementia. By CDR-SOB, with a proposed translation[25], 59.5% scored 0 (CN), 34.7% ranged 0.5–4.0 (MCI/SCC), and 5.8% ranged 4.5–9.0 (mild dementia). Missing data were minimal (≤ 5% across all variables, none for imaging, demographics, CDR-SOB, or diagnosis), with the exception of CSF measures, which had approximately 23% missingness. No significant group differences for demographic, cognitive measures (e.g., CDR-SOB p = 0.695, MMSE p = 0.753, ADAS-COG p = 0.828), or fluid biomarkers were observed across training, validation, and test sets (see Table 1).

**Table 1.**
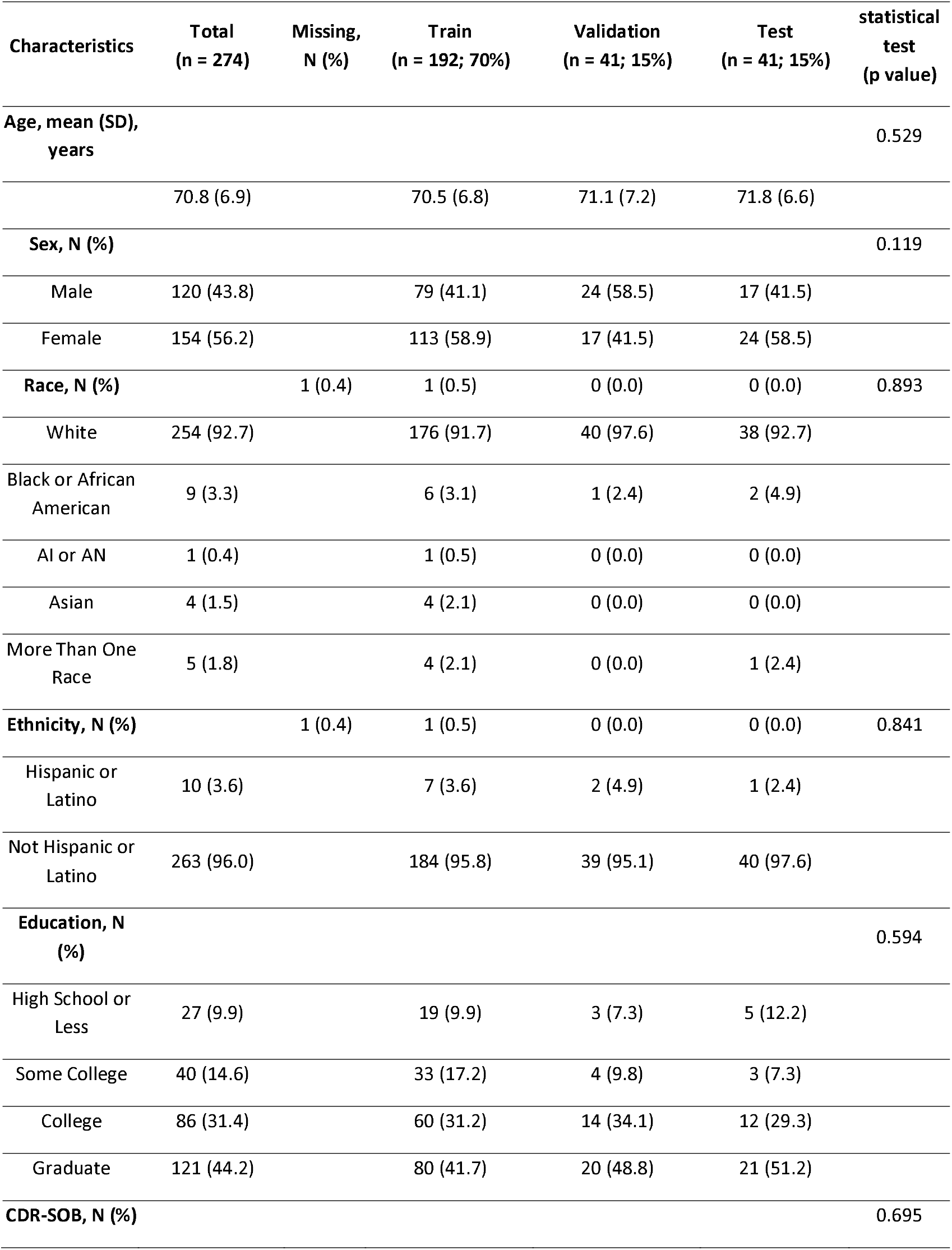
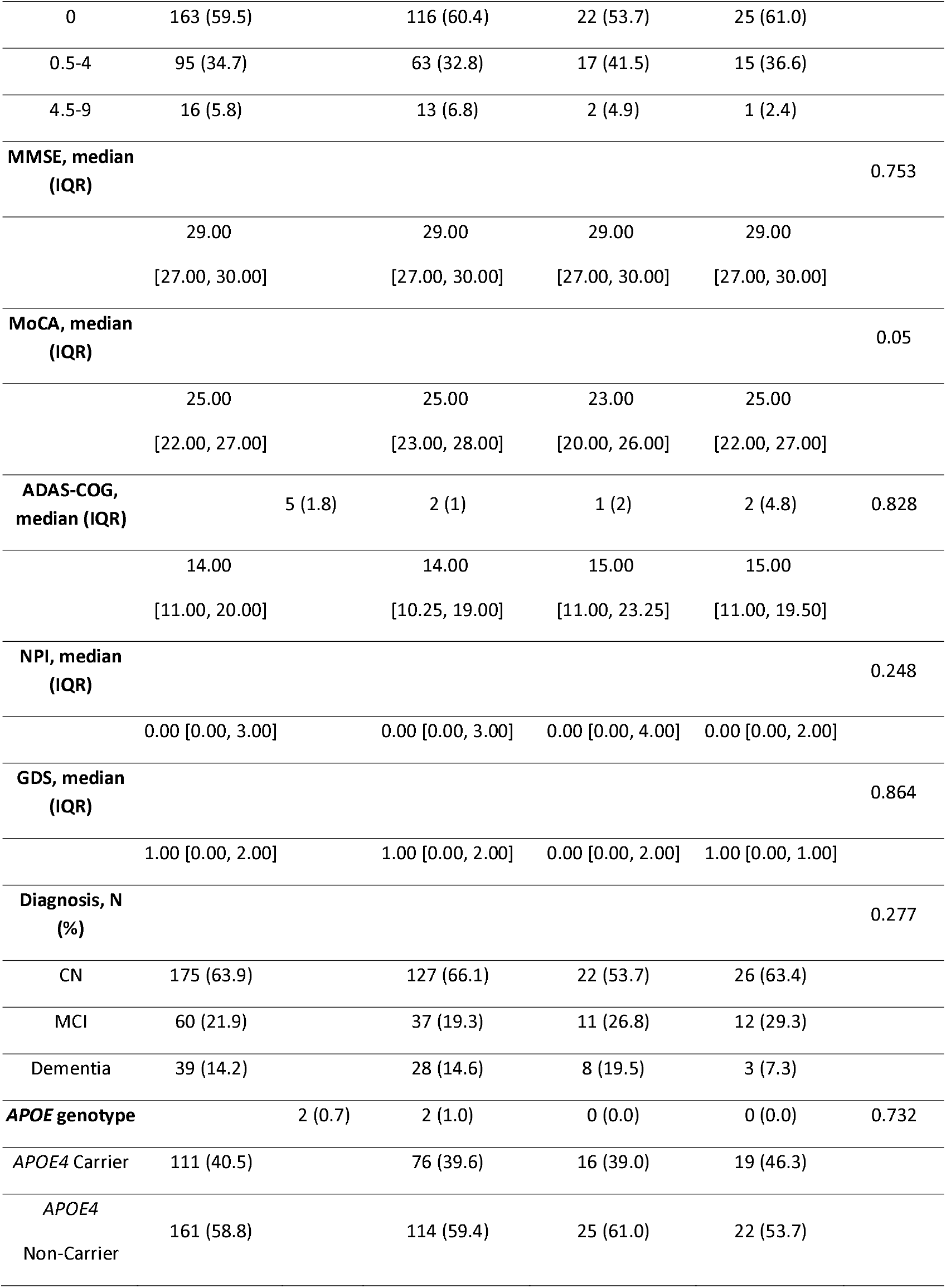
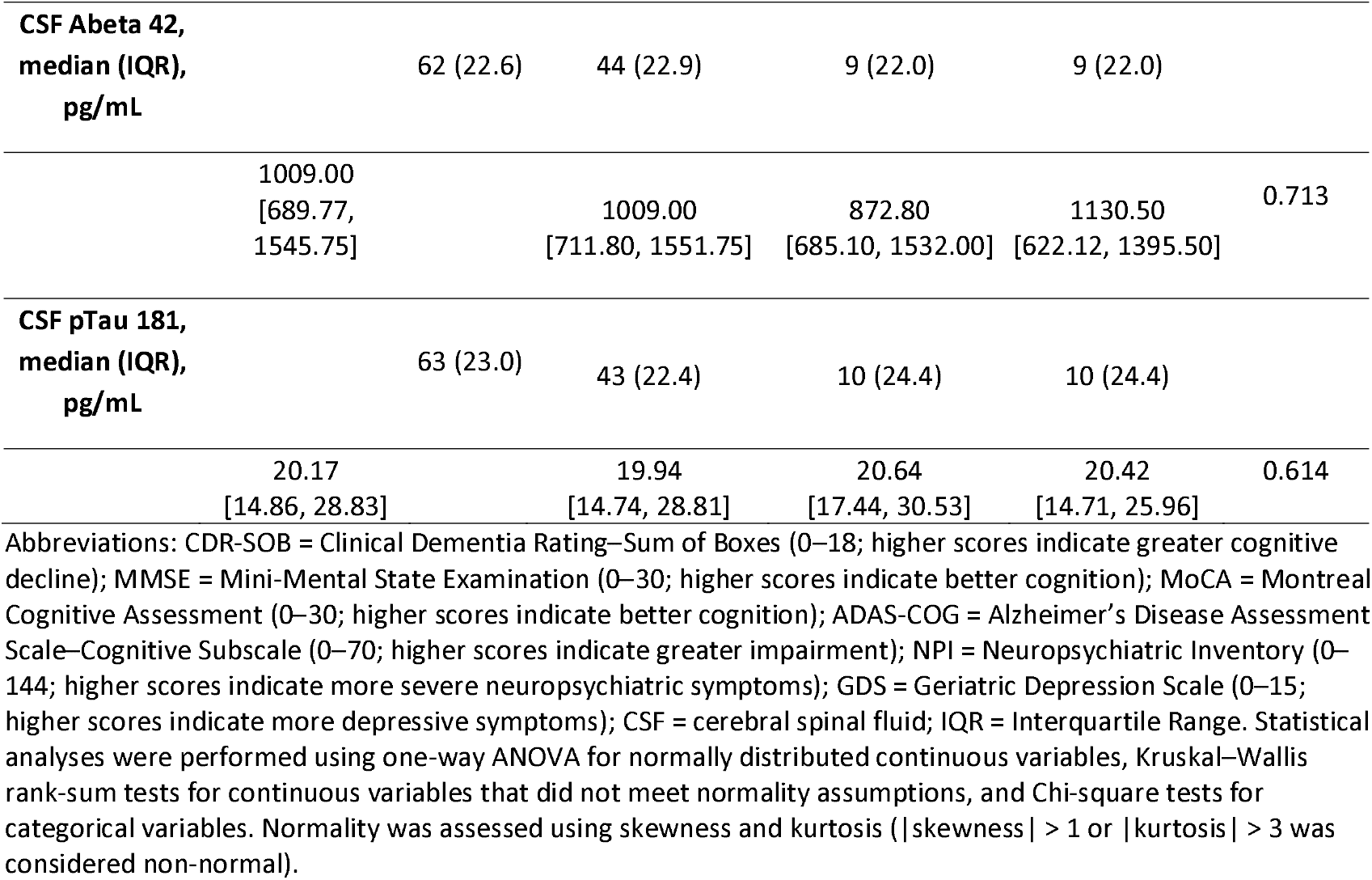
Summary statistics of the study population.

### 3.2 Multimodal components demonstrate model robustness

The SBF model achieved satisfactory performance, with a Pearson correlation of 0.21 between predicted and observed CDR-SOB scores in the independent test set (Figure 2A shows the prediction weight distribution across all 50 components; Figure 2B shows modality weight distributions within each latent component). After FDR multiple comparison, most of the components (92%; 46/50) showed a significant correlation with the target variable (i.e., CDR-SOB; Figure 2C), which further supports the robustness of our model prediction.

**Figure 2.**
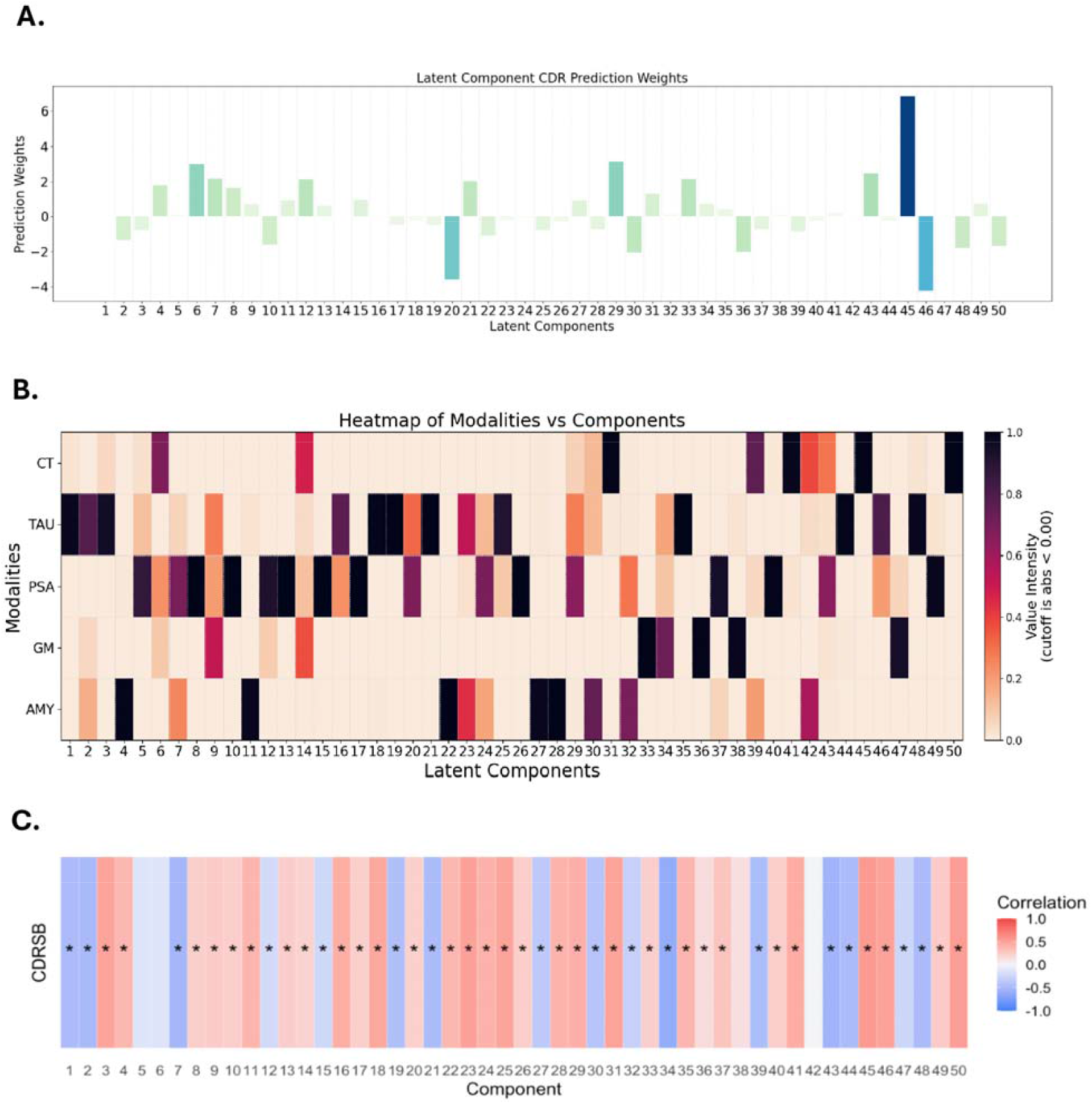
Weight distributions and component–CDR associations from SuperBigFLICA. A. Prediction weight distribution across 50 latent components. B. Modality weight distribution (unthresholded) showing the dominant modalities of each uni- or multi-modal component. C. Pearson correlations between component loadings and CDR-SOB show nearly all components were linked to CDR-SOB. Asterisks denote significance after Benjamini–Hochberg (BH) correction. Values indicate correlation coefficients; red reflects positive correlations; blue reflects negative correlations.

This investigation aimed to apply SBF to investigate the A-T-N framework for AD, specifically for identifying multi-modal brain patterns that are predictive of clinical diagnosis and *APOE4* carrier status. SBF predicts continuous variables, so CDR-SOB, which is related to both diagnostic and *APOE4* status, was used as the continuous target variable. Although diagnostic/*APOE4* status were not included in the training stage, SBF is effective for generating a representation space that has high predictive power for variables that are related to the training target[23].

### 3.3 SBF latent loadings outperform baseline methods for diagnostic prediction

The SBF loadings-based model outperformed all baseline methods—including prediction from demographics, single-modality PCA/ICA, and naïve-fusion PCA/ICA—achieving the highest macro-average AUROC 95% CI (0.80 [0.60, 0.92]), accuracy (0.84 [0.76, 0.90]), balanced accuracy (0.81 [0.72–0.88]), precision (0.91 [0.83–0.96]), and F1 score (0.88 [0.81–0.93]) (Table 2A). While baselines showed uneven sensitivity–specificity trade-offs, SBF consistently provided balanced gains across metrics (Table 2A, Table S1). Pairwise ROC analyses indicated strong performance for dementia discrimination (AUROC = 0.95 [0.82, 1] for CN vs. mild dementia, 0.89 [0.54, 1] for MCI vs. mild dementia), with weaker separation of CN vs. MCI (AUROC = 0.57 [0.46, 0.77]) (Figure 3A, B, D; Figure S1). AUPRC exhibited the same relative performance profile (Figures S1, S2; Tables 2A, S1), confirming that the observed discriminative ranking was not driven by test set imbalance. Note that since the highest CDR-SOB score among participants corresponds to the range categorized as mild dementia in the proposed translation[25], we specify the diagnosis of “dementia” as “mild dementia” for clarity.

**Table 2.**
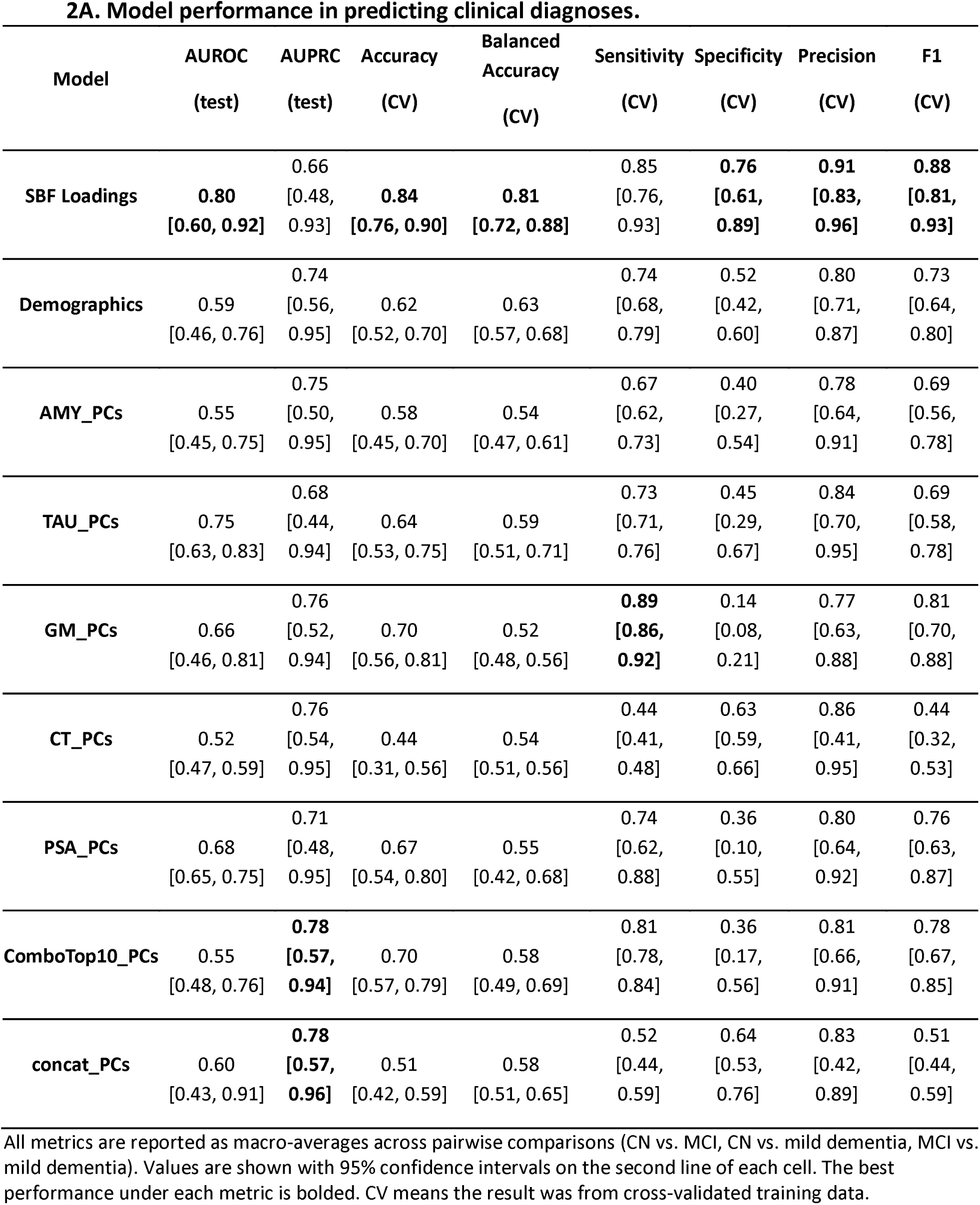
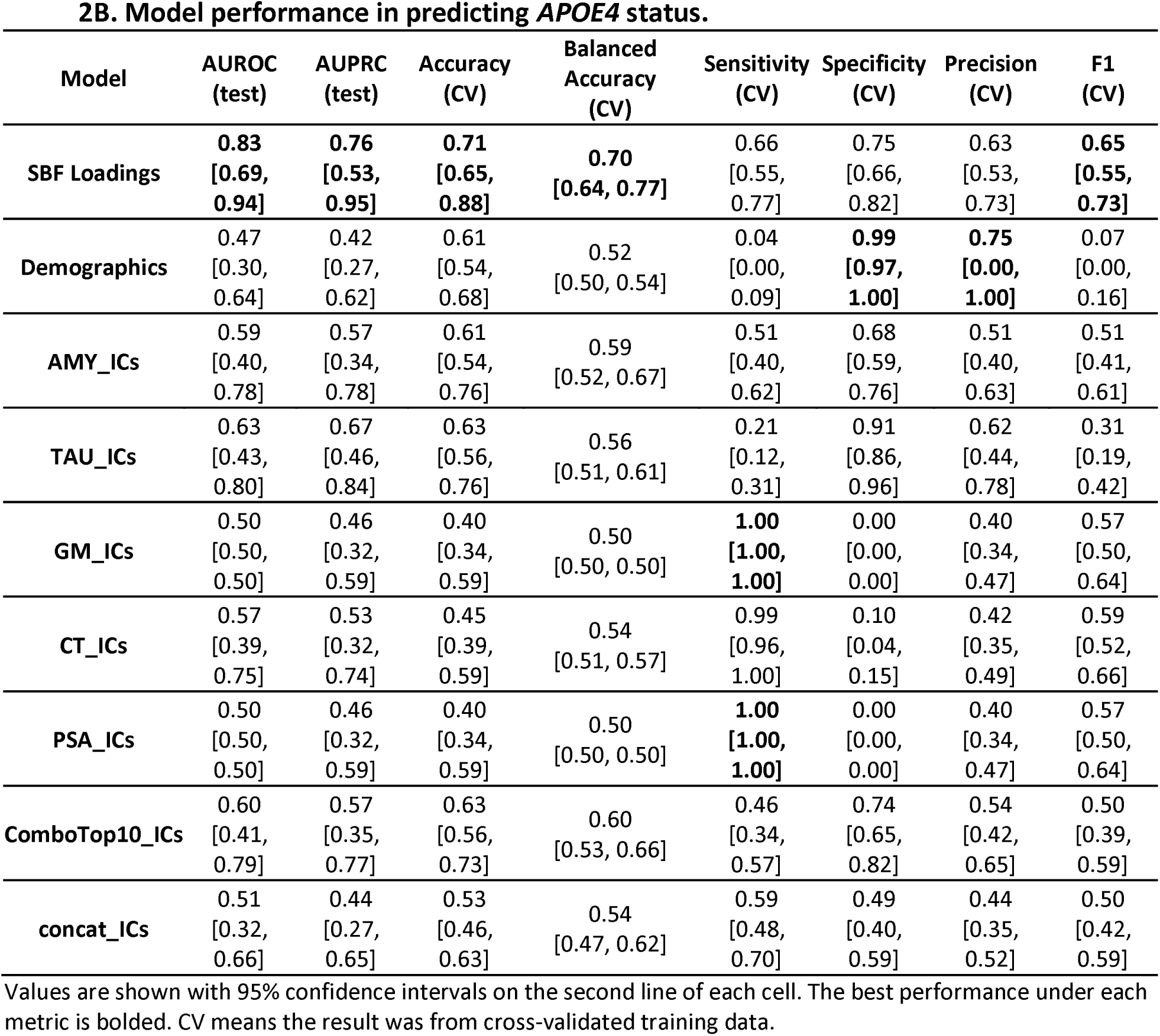
Model performance of SBF loadings–based models and baselines.

**Figure 3.**
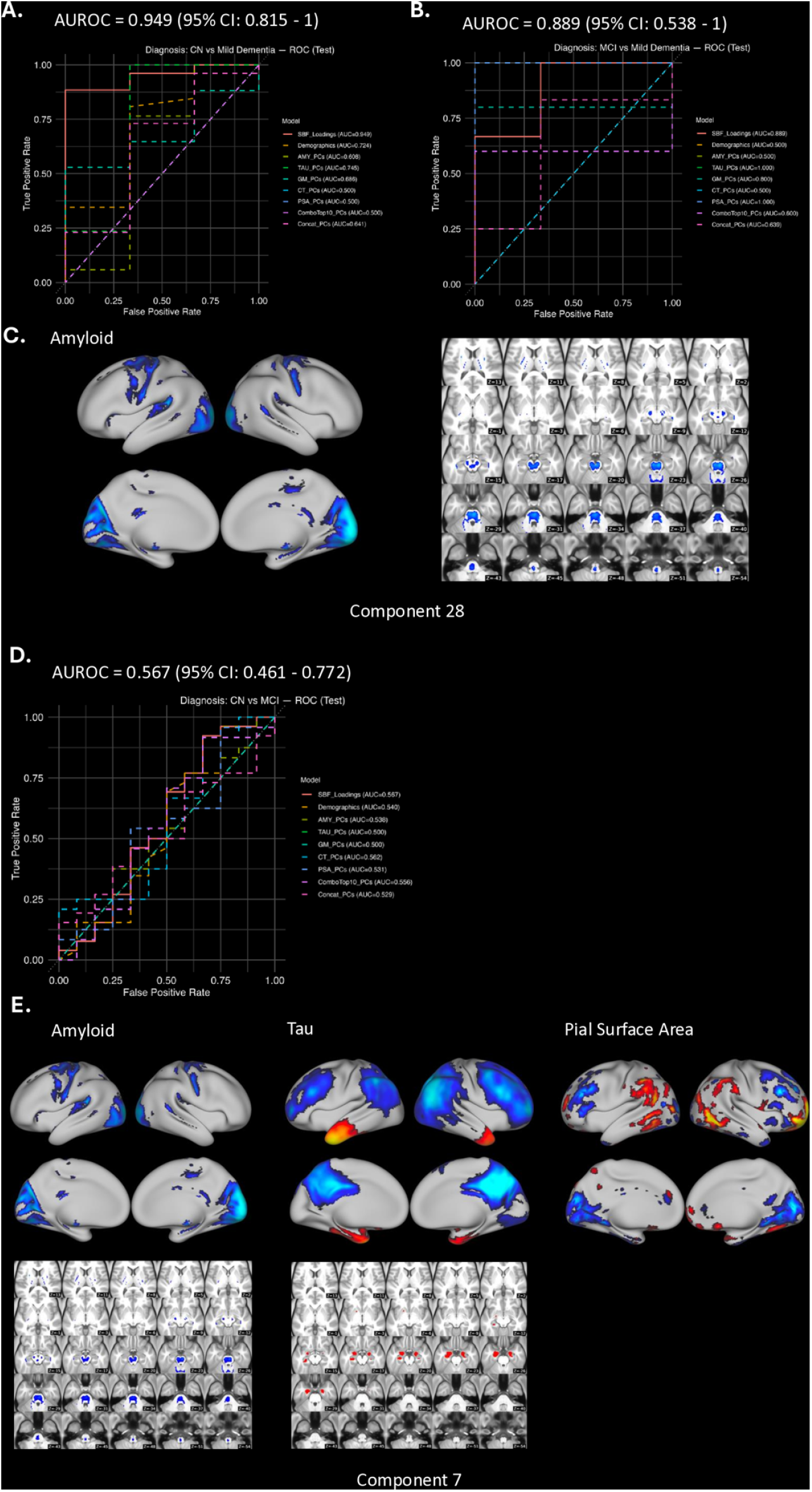
Model performance in predicting clinical diagnoses. A. ROC curve for CN vs. mild dementia. B. ROC curve for MCI vs. mild dementia. C. Spatial map of Component 28, the most predictive component for both CN vs. dementia and MCI vs. dementia, reflecting an amyloid-dominant pattern that represents low amyloid in these areas in CN and increasing amyloid with progression to MCI and early dementia. Left: cortical view. Right: subcortical view. D. ROC curve for CN vs. MCI. E. Spatial map of Component 7, the most predictive latent component for CN vs. MCI, reflecting a multimodal A-T-N pattern. This pattern links the low amyloid pattern in CN (seen in component 28) to normal aging-related tau and PSA patterns that weakens with progression to MCI and early dementia. This may result in uncoupling of amyloid with normal aging tau and PSA patterns as amyloid burden increases, ultimately leading to the amyloid pattern (component 28) best discriminating CN from MCI and early dementia. Upper row (left to right): cortical view for amyloid, tau, and pial surface area. Lower row (left to right): subcortical view for amyloid and tau. In (A, B, D), the AUROC listed on top of the legend (red line) corresponds to the SBF Loadings based model.

For mild dementia vs. CN and mild dementia vs. MCI, the most important feature was Component 28 (Figure 3C), a uni-modal amyloid-driven pattern spanning occipital, parietal, brainstem, and cerebellar regions, reflecting a relative preservation of amyloid burden in these areas as cognitive decline progresses to mild dementia. This is consistent with strong positive association between Component 28 loadings and CDR-SOB (Figure 2C).

For CN vs. MCI, the most important features were captured in Component 7 (Figure 3E), an A-T-N tri-modal pattern whose subject loadings were negatively associated with CDR-SOB (Figure 2C), suggesting a CN pattern that gets progressively weaker in MCI and mild dementia. This pattern is characterized by: low amyloid in occipital, parietal, brainstem, and cerebellar regions, resembling the amyloid pattern observed in Component 28, that covaries with higher tau in the anterior temporal lobe and temporal pole, and lower tau in inferior parietal, precuneus, posterior cingulate, fusiform, and temporal gyri. These molecular patterns were linked to lower surface area of orbitofrontal, inferior parietal, and lingual regions, that covaries with greater surface area of inferior frontal, temporal, anterior cingulate, lateral occipital, and supramarginal areas. This pattern of low amyloid burden but increasing tau in temporal areas, shrinkage of the cortical sheet in association areas and expansion in occipital and limbic areas, appears to represent a pattern of preserved cognitive function that weakens with increasing cognitive impairments (negative association between CDR-SOB and the subject loadings).

### 3.4 SBF latent loadings outperform baselines in *APOE4* prediction, driven by an amyloid pattern

The SBF loadings-based model achieved the highest performance in predicting *APOE4* carrier status (AUROC = 0.83 [0.69–0.94]; AUPRC = 0.76 [0.53–0.95]), exceeding demographics, single-modality, and naïve PCA/ICA fusion baselines (Figure 4A, 4B; Figure S3). Balanced accuracy (0.70 [0.64–0.77]) and F1 (0.65 [0.55–0.73]) also surpassed all comparators, indicating consistently stronger discrimination (Table 2B, Table S2). Similar to prediction of mild dementia vs. CN and mild dementia vs. MCI, the most predictive feature was again Component 28, an amyloid-driven pattern with prominent occipital and parietal involvement.

**Figure 4.**
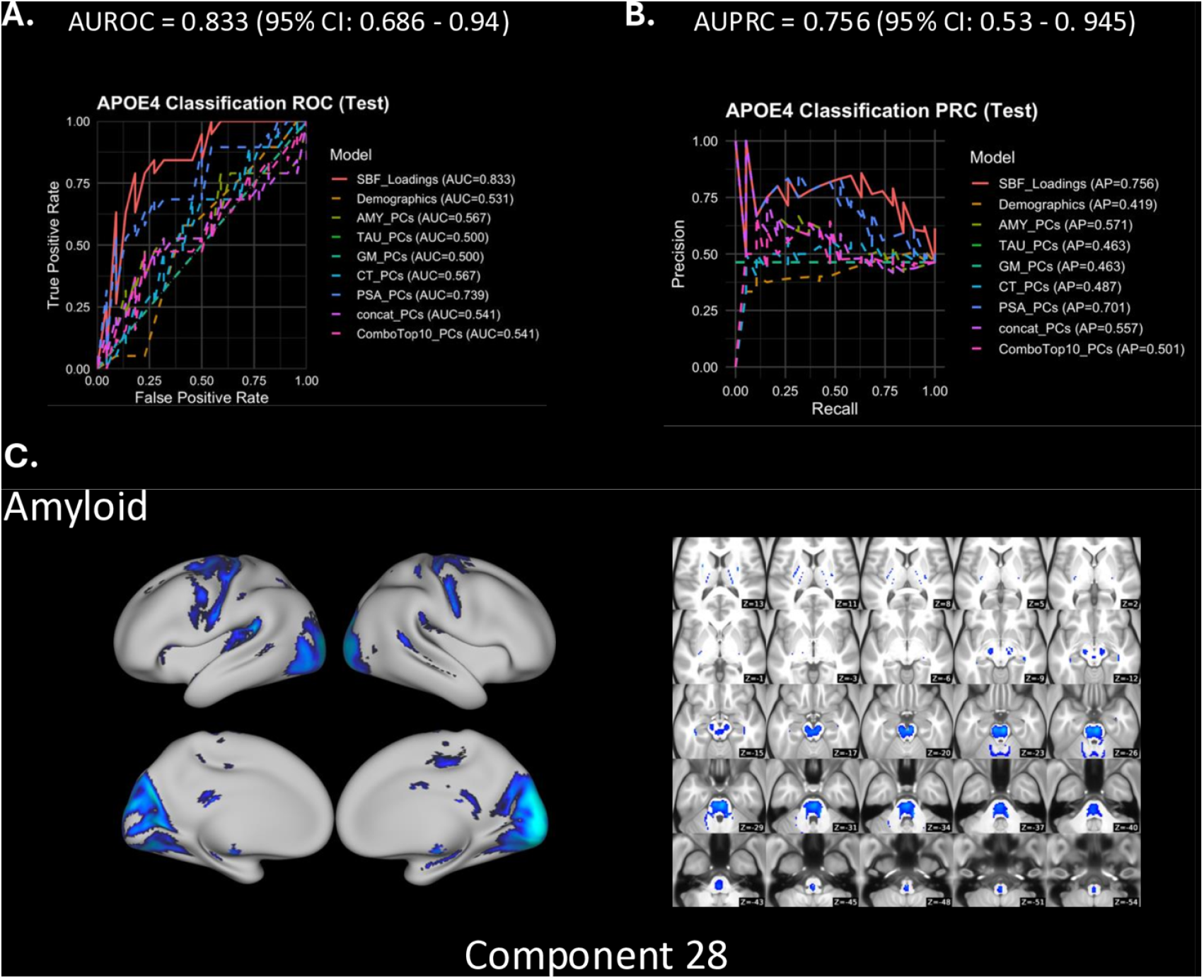
Model performance in predicting APOE4 status. A. ROC curve for *APOE4* carrier vs. non-carrier. B. PRC curve for *APOE4* carrier vs. non-carrier C. Spatial map of Component 28, the most predictive component for *APOE4* carrier vs. non-carrier, reflecting the same amyloid-dominant pattern that best discriminated CN and MCI from early dementia. Left: cortical view. Right: subcortical view. In (A, B), the AUROC and AUPRC listed on top of the legend (red line) corresponds to the SBF Loadings based model.

### 3.5 Component loadings capture distinct CSF biomarker associations

Among the 192 training participants, 148 (77.1%) had CSF AD biomarkers available for correlation analyses. Loadings for Component 7 showed a positive correlation with CSF Aβ42 (r = 0.4, p < .001), in which higher loadings (e.g., less amyloid deposition in the brain) were associated with higher CSF Aβ42, which is consistent with the interpretation that if Aβ42 is retained in CSF then there is lower Aβ42 in the brain, and a negative correlation with CSF pTau181 (r = −0.55, p < .001), reflecting lower tau burden. In contrast, loadings on Component 28 showed the opposite pattern: negative correlation with CSF Aβ42 (r = −0.53, p < .001) and positive correlation with CSF pTau181 (r = 0.59, p < .001), consistent with higher amyloid and tau burden in the brain.

## Discussion

In this work, we applied SuperBigFLICA (SBF), a semi-supervised multimodal data fusion framework, to investigate the A-T-N framework for AD continuum. Unlike conventional uni-modal or naïve-fusion approaches, SBF identifies latent components that simultaneously maximizes data reconstruction and prediction accuracy of non-imaging phenotypes. This enables joint statistical integration of brain imaging with cognitive and behavioral measures, enhancing the interpretability of the resulting multimodal patterns. Using ADNI-3 data across five imaging modalities (amyloid PET, tau PET, gray matter density, cortical thickness, and pial surface area), we demonstrated that SBF can capture meaningful A-T-N brain–behavior relationships with cognitive decline (CDR-SOB) used as a training target variable (r = 0.21). More than 90% of latent components were significantly associated with the target variable, underscoring that the model identified disease-relevant multimodal signals. We further applied the component loadings to predict additional outcomes, including clinical diagnoses and *APOE4* carrier status. In both cases, the SBF-derived models outperformed single-modality and unsupervised naïve fusion baselines (PCA/ICA), showing that SBF’s latent imaging components captured integrated disease-related variation that extends beyond the training target. These findings demonstrate that SBF not only improves prediction of its training target but also uncovers a high-dimensional representational space that generalizes to related phenotypes.

SBF maintains interpretability because the projection from subject loadings back to modality-specific spatial maps is fully linear, rather than routed through a black-box predictive model. This interpretability, in turn, enables explainability, meaning the spatial patterns supporting a prediction can be examined directly instead of remaining opaque latent representations. As a result, SBF does not simply predict a neuropsychological score — it surfaces the spatial patterns associated with that prediction and not other sources of intersubject variability, enabling principled inference about the dominant neuroimaging signature driving the outcome, whether unimodal or multimodal. This is an advantage over existing data fusion frameworks that can identify multimodal patterns, but these patterns may reflect many different sources of intersubject variability, requiring post hoc analyses to identify disease patterns.

With CDR-SOB as a proxy target variable, separation of CN from MCI was best captured by a multimodal covariance pattern spanning A-T-N modalities (Component 7). Weaker classification performance–regardless of the features used—highlights the challenge of detecting early cognitive decline. In line with this, clinicians reported low confidence in identifying MCI in real-world practice[42,43], and variability in neuroimaging biomarkers causes considerable overlap with the healthy population, limiting their ability to reliably discriminate CN from MCI[44]. Considering the relationships between loadings, CDR-SOB, and prediction performance, the A-T-N pattern is likely a normal aging cognitive decline pattern that is weaker in MCI.

In contrast, distinguishing mild dementia from CN or MCI—especially mild dementia against MCI, where clinical distinctions remain challenging—was dominated by a single-modality amyloid pattern of deposition in lower-order sensory areas (Component 28)[45] that achieved high predictive accuracy for both tasks. These visual and motor areas are the first to develop in a hierarchical sensorimotor-to-association axis of cortical organization[45] and align with functional gradients reported to be altered in normal aging and AD[46]). Interestingly, the patterns of low amyloid and tau in component 7 reflect these two dichotomous axes, with low amyloid in sensorimotor areas and low tau in association areas. CSF analyses revealed that Component 7 loadings were associated with lower amyloid and tau burden, aligning with normal aging. In contrast, Component 28 loadings were linked to greater amyloid and tau burden (measured by CSF), aligning with cognitive decline and disease progression. Component 7 additionally reflected tau deposition in medial temporal nodes, suggesting that tau’s contribution is more characteristic of aging- and early-stage-related cognitive decline, whereas amyloid-driven deposition seen in component 28 is characteristic of later MCI and mild dementia disease stages not linked to tau deposition through SBF.

Importantly, beyond molecular pathology, pial surface area (PSA) of the cortical sheet was linked to low amyloid and tau in component 7. The comparable performance of PSA PCs to tau PCs for discriminating MCI vs mild dementia suggests that PSA may be a sensitive biomarker for early-stage detection. Nevertheless, PSA remains an underexplored structural marker relative to cortical thickness. Our findings highlight its potential relevance and motivate deeper investigation moving forward. Finally, the same amyloid-driven sensory pattern (Component 28) that best distinguished mild dementia from CN and MCI also emerged as most predictive of *APOE4* carrier status. This reinforces this component’s role as a convergent marker of both genetic risk and clinical progression.

Looking ahead, SBF’s interpretability makes it well-suited for integrating additional modalities into a unified and biologically coherent representation space. Incorporating additional modalities, such as functional and structural connectivity, may improve predictive performance and mechanistic insight while maintaining SBF’s interpretability. Some limitations of the present study are its cross-sectional design; applying the framework to longitudinal data could yield insights into temporal disease dynamics. A second limitation is the modest sample size for prediction. However, SBF exploits joint information among modalities and within participants and embeds prediction directly into the decomposition to identify multimodal maps that specifically focus on explaining variance in the target variable. Combined with rigorous methods for cross-validation and inclusion of a small independent test sample greatly improves the rigor of our findings.

## Conclusions

Applying SuperBigFLICA (SBF) to investigate the A-T-N framework for AD demonstrates that semi-supervised multimodal fusion can capture continuous variation across individuals (e.g., cognitive decline), while also uncovering integrative brain patterns linked to diagnosis and genetic risk. This highlights SBF’s methodological value for heterogeneous, multimodal disease settings, where interpretability is integral rather than sacrificed for predictive performance. Moving forward, such approaches hold promise for advancing precision staging and guiding targeted interventions in dementia research and care.

## CODE AVAILABILITY

Upon acceptance, all code used for multimodal data fusion and model training will be available at: https://github.com/ANSR-laboratory/SuperBigFLICA_McL. A detailed model card describing the model architecture, inputs/outputs, evaluation, and intended use will be found at: https://github.com/ANSR-laboratory/SuperBigFLICA_McL/blob/main/model_cards/sbf_cdrsob_adni3_MODEL_CARD.md.

## DATA AVAILABILITY

Neuroimaging and clinical data were obtained from the Alzheimer’s Disease Neuroimaging Initiative (ADNI; http://adni.loni.usc.edu) and can be accessed upon approval of a data use application.

## ACKNOWLEDGEMENTS

Data collection and sharing for this project was funded by the Alzheimer’s Disease. Neuroimaging Initiative (ADNI) (National Institutes of Health Grant U01 AG024904) and DOD ADNI (Department of Defense award number W81XWH-12-2-0012). ADNI is funded by the National Institute on Aging, the National Institute of Biomedical Imaging and Bioengineering, and through generous contributions from the following: AbbVie, Alzheimer’s Association; Alzheimer’s Drug Discovery Foundation; Araclon Biotech; BioClinica, Inc.; Biogen; Bristol-Myers Squibb Company; CereSpir, Inc.; Cogstate; Eisai Inc.; Elan Pharmaceuticals, Inc.; Eli Lilly and Company; EuroImmun; F. Hoffmann-La Roche Ltd and its affiliated company Genentech, Inc.; Fujirebio; GE Healthcare; IXICO Ltd.; Janssen Alzheimer Immunotherapy Research & Development, LLC.; Johnson & Johnson Pharmaceutical Research & Development LLC.; Lumosity; Lundbeck; Merck & Co., Inc.; Meso Scale Diagnostics, LLC.; NeuroRx Research; Neurotrack Technologies; Novartis Pharmaceuticals Corporation; Pfizer Inc.; Piramal Imaging; Servier; Takeda Pharmaceutical Company; and Transition Therapeutics. The Canadian Institutes of Health Research is providing funds to support ADNI clinical sites in Canada. Private sector contributions are facilitated by the Foundation for the National Institutes of Health (www.fnih.org). The grantee organization is the Northern California Institute for Research and Education, and the study is coordinated by the Alzheimer’s Therapeutic Research Institute at the University of Southern California. ADNI data are disseminated by the Laboratory for Neuro Imaging at the University of Southern California.

## CONSENT STATEMENT

All participants provided written informed consent, and each ADNI site obtained local institutional review board approval for study procedures.

