## supplementary_materials1 for "Investigating the Amyloid-Tau-Neurodegeneration Framework in Alzheimer’s Disease Using Semi-Supervised Multimodal Imaging Data Fusion"

**Supplements**

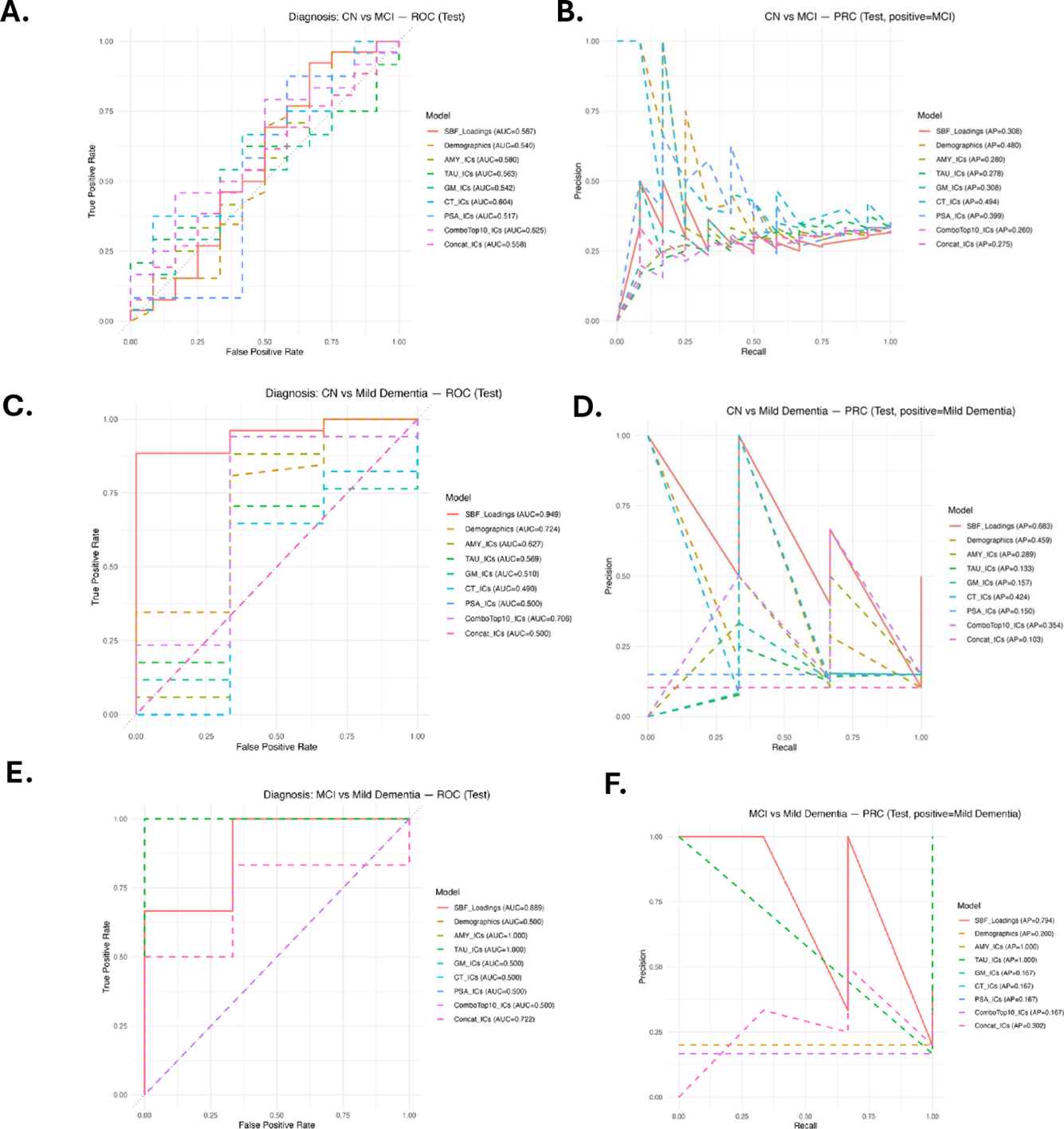

**Figure S1. Model performance in predicting clinical diagnoses, comparing SBF loadings–based models, demographic baseline, and ICA baselines.**

A. ROC curve for CN vs. MCI. B. PRC curve for CN vs. MCI. C. ROC curve for CN vs. dementia. D. PRC curve for CN vs. dementia. E. ROC curve for MCI vs. dementia. F. PRC curve for MCI vs. dementia. The analyses indicated that, compared with all baselines, the model based on SBF loadings strong performance for dementia discrimination for CN vs. mild dementia and MCI vs. mild dementia, with weaker separation of CN vs. MCI. AUPRC exhibited the same relative performance profile, confirming that the observed discriminative ranking was not driven by test set imbalance.

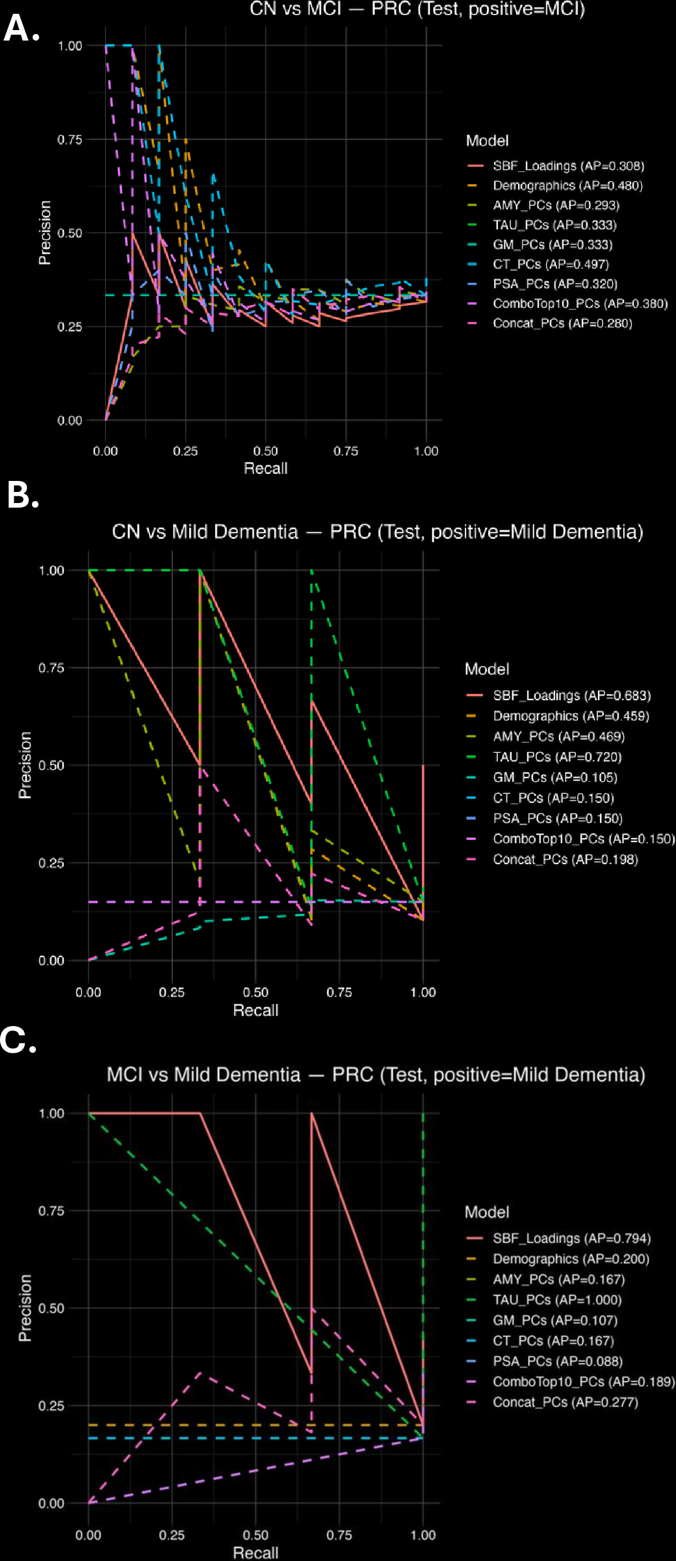

**Figure S2. Model performance in predicting clinical diagnoses.**

A.PRC curve for CN vs. MCI. B. PRC curve for CN vs. dementia. C. PRC curve for MCI vs. dementia. The results indicated that, compared with all baselines, the model based on SBF loadings had strong performance for dementia discrimination for CN vs. mild dementia, MCI vs. mild dementia, with weaker separation of CN vs. MCI.

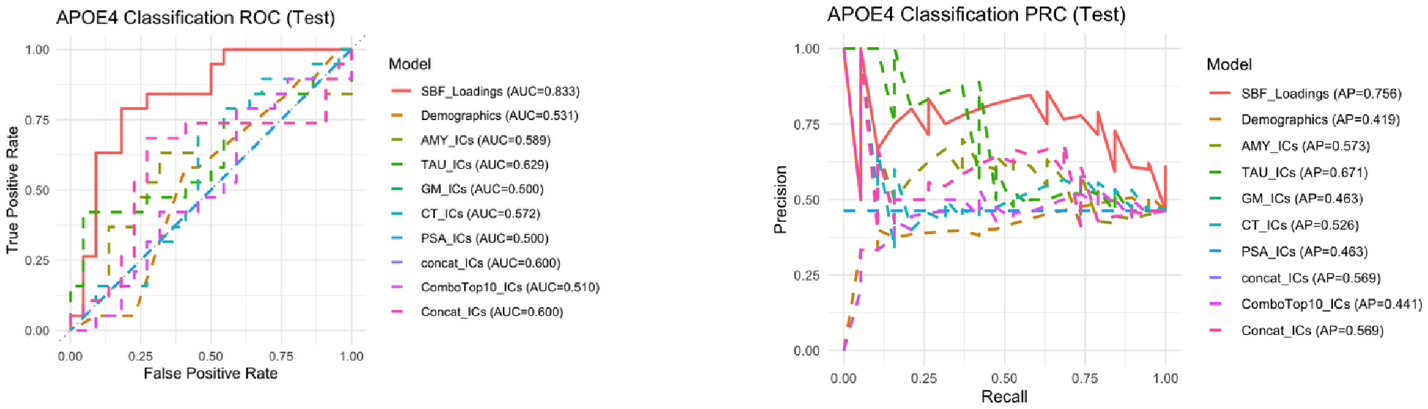

**Figure S3.** **Model performance in predicting *APOE4* status, comparing SBF loadings–based models, demographic baseline, and ICA baselines.**

A. ROC curve for *APOE4* carrier vs. non-carrier. B. PRC curve for *APOE4* carrier vs. non-carrier. The results showed the model based on SBF loadings had stronger performance for *APOE4* carrier/non-carrier discrimination than all baselines.

**Table S1. Model performance in predicting clinical diagnoses, comparing SBF loadings–based models, demographic baseline, and ICA baselines.**

| **Model** | **AUROC**  **(test)** | **AUPRC**  **(test)** | **Accuracy**  **(CV)** | **Balanced Accuracy**  **(CV)** | **Sensitivity**  **(CV)** | **Specificity**  **(CV)** | **Precision**  **(CV)** | **F1**  **(CV)** |
| --- | --- | --- | --- | --- | --- | --- | --- | --- |
| **SBF Loadings** | **0.80 [0.60, 0.92]** | 0.66 [0.48, 0.93] | **0.84 [0.76, 0.90]** | **0.81 [0.72, 0.88]** | **0.85 [0.76, 0.93]** | **0.76 [0.61, 0.89]** | **0.91 [0.83, 0.96]** | **0.88 [0.81, 0.93]** |
| **Demographics** | 0.59 [0.46, 0.76] | 0.74 [0.56, 0.95] | 0.62 [0.52, 0.70] | 0.63 [0.57, 0.68] | 0.74 [0.68, 0.79] | 0.52 [0.42, 0.60] | 0.80 [0.71, 0.87] | 0.73 [0.64, 0.80] |
| **AMY_ICs** | 0.74 [0.62, 0.92] | 0.68 [0.44, 0.94] | 0.41 [0.27, 0.54] | 0.58 [0.50, 0.67] | 0.24 [0.13, 0.37] | 0.93 [0.84, 1.00] | 0.93 [0.52, 1.00] | 0.37 [0.20, 0.53] |
| **TAU_ICs** | 0.71 [0.63, 0.88] | 0.73 [0.48, 0.95] | 0.53 [0.39, 0.65] | 0.58 [0.45, 0.73] | 0.52 [0.41, 0.61] | 0.64 [0.41, 0.94] | 0.83 [0.67, 0.97] | 0.59 [0.46, 0.70] |
| **GM_ICs** | 0.52 [0.45, 0.73] | **0.78 [0.54, 0.95]** | 0.55 [0.42, 0.67] | 0.52 [0.48, 0.55] | 0.63 [0.59, 0.66] | 0.41 [0.35, 0.47] | 0.84 [0.73, 0.92] | 0.58 [0.47, 0.65] |
| **CT_ICs** | 0.53 [0.45, 0.76] | 0.74 [0.51, 0.94] | 0.52 [0.39, 0.63] | 0.52 [0.49, 0.56] | 0.59 [0.56, 0.63] | 0.45 [0.39, 0.51] | 0.83 [0.72, 0.92] | 0.55 [0.45, 0.63] |
| **PSA_ICs** | 0.51 [0.48, 0.59] | 0.77 [0.54, 0.95] | 0.65 [0.52, 0.76] | 0.53 [0.50, 0.55] | 0.79 [0.76, 0.82] | 0.26 [0.22, 0.31] | 0.79 [0.65, 0.90] | 0.74 [0.63, 0.82] |
| **ComboTop10_ICs** | 0.61 [0.46, 0.77] | 0.73 [0.48, 0.94] | 0.41 [0.28, 0.52] | 0.52 [0.50, 0.54] | 0.40 [0.37, 0.43] | 0.65 [0.62, 0.67] | 0.87 [0.41, 0.96] | 0.38 [0.27, 0.46] |
| **concat_ICs** | 0.59 [0.46, 0.75] | 0.77 [0.58, 0.93] | 0.70 [0.62, 0.78] | 0.58 [0.52, 0.65] | 0.83 [0.76, 0.89] | 0.34 [0.23, 0.46] | 0.77 [0.68, 0.86] | 0.79 [0.71, 0.85] |

All metrics are reported as macro-averages across pairwise comparisons (CN vs. MCI, CN vs. dementia, MCI vs. dementia). Values are shown with 95% confidence intervals on the second line of each cell. The best performance under each metric is bolded. CV means the result was from cross-validated training data.

**Table S2. Model performance in predicting *APOE4* status, comparing SBF loadings–based models, demographic baseline, and ICA baselines.**

| **Model** | **AUROC**  **(test)** | **AUPRC**  **(test)** | **Accuracy**  **(CV)** | **Balanced Accuracy**  **(CV)** | **Sensitivity**  **(CV)** | **Specificity**  **(CV)** | **Precision**  **(CV)** | **F1**  **(CV)** |
| --- | --- | --- | --- | --- | --- | --- | --- | --- |
| **SBF Loadings** | **0.83 [0.69, 0.94]** | **0.76 [0.53, 0.95]** | **0.71 [0.65, 0.88]** | **0.70 [0.64, 0.77]** | 0.66 [0.55, 0.77] | 0.75 [0.66, 0.82] | 0.63 [0.53, 0.73] | **0.65 [0.55, 0.73]** |
| **Demographics** | 0.47 [0.30, 0.64] | 0.42 [0.27, 0.62] | 0.61 [0.54, 0.68] | 0.52 [0.50, 0.54] | 0.04 [0.00, 0.09] | **0.99 [0.97, 1.00]** | **0.75 [0.00, 1.00]** | 0.07 [0.00, 0.16] |
| **AMY_ICs** | 0.59 [0.40, 0.78] | 0.57 [0.34, 0.78] | 0.61 [0.54, 0.76] | 0.59 [0.52, 0.67] | 0.51 [0.40, 0.62] | 0.68 [0.59, 0.76] | 0.51 [0.40, 0.63] | 0.51 [0.41, 0.61] |
| **TAU_ICs** | 0.63 [0.43, 0.80] | 0.67 [0.46, 0.84] | 0.63 [0.56, 0.76] | 0.56 [0.51, 0.61] | 0.21 [0.12, 0.31] | 0.91 [0.86, 0.96] | 0.62 [0.44, 0.78] | 0.31 [0.19, 0.42] |
| **GM_ICs** | 0.50 [0.50, 0.50] | 0.46 [0.32, 0.59] | 0.40 [0.34, 0.59] | 0.50 [0.50, 0.50] | **1.00 [1.00, 1.00]** | 0.00 [0.00, 0.00] | 0.40 [0.34, 0.47] | 0.57 [0.50, 0.64] |
| **CT_ICs** | 0.57 [0.39, 0.75] | 0.53 [0.32, 0.74] | 0.45 [0.39, 0.59] | 0.54 [0.51, 0.57] | 0.99 [0.96, 1.00] | 0.10 [0.04, 0.15] | 0.42 [0.35, 0.49] | 0.59 [0.52, 0.66] |
| **PSA_ICs** | 0.50 [0.50, 0.50] | 0.46 [0.32, 0.59] | 0.40 [0.34, 0.59] | 0.50 [0.50, 0.50] | **1.00 [1.00, 1.00]** | 0.00 [0.00, 0.00] | 0.40 [0.34, 0.47] | 0.57 [0.50, 0.64] |
| **ComboTop10_ICs** | 0.60 [0.41, 0.79] | 0.57 [0.35, 0.77] | 0.63 [0.56, 0.73] | 0.60 [0.53, 0.66] | 0.46 [0.34, 0.57] | 0.74 [0.65, 0.82] | 0.54 [0.42, 0.65] | 0.50 [0.39, 0.59] |
| **concat_ICs** | 0.51 [0.32, 0.66] | 0.44 [0.27, 0.65] | 0.53 [0.46, 0.63] | 0.54 [0.47, 0.62] | 0.59 [0.48, 0.70] | 0.49 [0.40, 0.59] | 0.44 [0.35, 0.52] | 0.50 [0.42, 0.59] |

Values are shown with 95% confidence intervals on the second line of each cell. The best performance under each metric is bolded. CV means the result was from cross-validated training data.
